# Assessment of menstrual hygiene practice and associated factor among High school female students in Harari Region, Eastern Ethiopia 2019

**DOI:** 10.1101/2020.03.16.20036913

**Authors:** Astawus Alemayehu, Abdi Ahmed, Maruf Abdalla

## Abstract

**Background:** Menstruation is a visible manifestation of cyclic uterine bleeding as a result of the interaction of different hormones. During menstruation, girls face gender problems. These are, early marriage, premature childbirth, higher infant mortality and potential vaginal infections resulting in infertility.

**Objective:** To assess the level of menstrual hygiene practice and associated factors among high school female students in Harari Region Eastern, Ethiopia, 2019.

**Study Design and Method:** A cross-sectional quantitative study was employed from April 02-05/2019. A minimum sample size of 301 Data was collected using a self-administered structured questionnaire. Descriptive and analytical facts were applied. Bivariate analysis and multivariate regression model were used. P-value <= 0.05 was declared as statistical significance

**Result:** According to study 168(55.8%) had good practiced the rest had no. students who have no pocket money from family(AOR 0.36:95% CI,0.309,0.989) was 64% less likely poor practiced than students who have permanent pocket money, students who have no educated father(AOR 0.39:95% CI, 0.180,0.872) was 61% less likely poor practiced than who have educated father and students who had not Freely discuss with parents(AOR 0.45:95% CI,0.22,0.903) was 55% less likely poor practiced than who had Freely discuss with parents.

**Conclusion:** Majority of female students in Harar region had good knowledge about menstrual hygiene practice. good menstrual hygiene practice was more among students who live in the urban than students who live in the rural area; students who have permanent pocket money from family than students who have no permanent pocket money from family.

## 1. Introduction

In 2011 globally, around fifty-two percent of the lady population in reproductive age, Most of these ladies menstruate every month for two and seven days. Girls usually start to menstruate throughout puberty or adolescence. MHM is the absorption menstrual blood onto smooth cloth which may be changed in privacy. Which consisting of the supply of cleaning soap and easy water, to clean reusable sanitary materials and body, in addition to a suitable place of disposal. [1, 2]

Globally 20% of girls miss school due to their monthly period, and one in ten will drop out altogether. 39% and 8%of girls use water however no soap for washing or cleaning their menstruation in India and Afghanistan respectively. Ladies Participation and performance became affected at school during menses. In various studies from Africa, the Middle East, and Asia, up to 95% of girls reported missing school because of their periods. In Malawi, 30% of girls don’t use the latrine during menstruation. There are 20% girls similarly observed in India communities and 11% and 60% ladies handiest trade their menstrual cloths as soon as a day in Ethiopia and India respectively. A similar study in Udupi Taluk, India showed that only 35.82% rural and 33.27% of urban participants had awareness about menses prior to menarche. [3-5]

A study in Mehalmeda northern Ethiopia indicates that all respondents use a menstrual sanitary pad, 70.9% use sanitary pad/napkins from market, 26.01% homemade cleaning cloths and 3.4% use other, 29.7% of the ladies reuse sanitary pad. A similar study in Ethiopia indicated that out of 455 ladies who had their menarche, 85.49 % of them did no longer trade menstrual absorb at school and the principal motives had been the absence of separate toilet for woman college students is 45.50%, worry other college students is 39.07%, lack of water assets is 18.77% and shortage of sanitary napkins/pads 15.17%. [6, 7]

In Nigeria 64.2% of the participants knew about menstruation and mother is an essential supply of information, while buddies and TV also contributed to their information. other similar study in Northwest Ethiopia shows, out of 423 study participants 29.8% of the respondents practiced good menstrual hygiene whereas 70.2% practiced poor menstrual hygiene. Knowledge regarding menstrual hygiene was positively associated with practice on menstrual hygiene. This finding was consistent with other studies. [8-10] The study aims to assess menstrual hygiene practice of girls’ in high school and identify associated factor, a result of this study is important to prevent hassle which linked to negative menstrual hygiene exercise. Due to its direct effect on –school absenteeism increased susceptibility to RTIs, infections of the perineum, elevated the potential risk of contracting blood-borne diseases consisting of HIV or HBV and gender discrepancy.

## 2. Study Design and Methods

### 2.1 Study areas and Design

The study was employed in Harari regional states, which is located in eastern part of Ethiopia. The regional states of Harari found 516 k/m away from the Addis Ababa city with an elevation of 1830 meters above the sea level. The total population of Harare regional state is estimated to be 232,000. From the total population, 62.6% live in urban and the rest 37.4 % live in rural areas. The regional state has six urban and three rural Districts (Woreda) and further the region is subdivided into 19 urban and 17 rural Keble’s. Majors ethnic groups within the region are Oromo, Amhara and Harari; and the main religions followed are Islam and Ethiopian Orthodox. According to Harari Educational Bureau 2011 E.C (2019 G.C) report, there are 8 governmental secondary schools and 9 non-governmental secondary schools. Total female and male students in both secondary schools are 7550, Only 1887 female students learn grades 9 and 10. A school-based quantitative cross-sectional study design was conducted from April 02 - 05/2019.

### 2.2 Sample size and sampling technique

The sample size turned into decided by using the usage of single population proportion formula with the assumptions of 95% confidence interval, a marginal error of (d = 5%), and through taking 29.8% considering the proportion of students who practiced good menstrual hygiene from studies done in the country, the sample size was 321 Since the source population was less than 10,000 populations the correction formula was used and considering 10 % non-response rate the final sample size becomes 301. The sample size was proportionally allocated accordingly, A simple random and systematic sampling was done to select girls on both governmental and private secondary school girls. Female students in regular programs who were starting menstruating at least one time before data collection and who are volunteer to participate in the study were included.

### 2.3. Study variables

The outcome variable in this look at turned into menstrual hygiene practice (poor practiced, good practiced). Independent variables include; socio-demographic variable; Sanitary material related variables (Availability, Affordability); School Environmental related variables (Presence of latrine, pad to manage period, continuous water supply, separate bathrooms for girls, private place to manage period at school); Knowledge& Source of information related variables.

### 2.4. Data assortment tools and procedures

The data or information have been gathered through a self-directed technique; questionnaire adapted from relevant literature of similar studies. It consists of basic socio-demographic characteristics, source of information or knowledge and awareness regarding menses, menstrual hygiene practice and associated factors and the questioner was prepared in the English version and was translated back to Amharic and Afan Oromo (local language) to check its consistency. To ascertain the quality of data, 5% of the total sample size pre-test questioner was done on Rainbow high schools female students and training were given to data gatherers and over lookers for one day on how to approach study subjects, on how to use the questionnaire and also Supervision was done on the spot by investigators and supervisors.

### 2.5 Data process and analysis

Before information entered into SPSS v.20 for analysis information was coded and checked for uniformity and completeness. Descriptive and analytical facts were applied, Also Bivariate analysis was used to examine the association between dependent and independent variables. All variables within the bivariate statistics P-value <=0.20 were moved into the multivariate regression model to identify associated factors with the menstrual hygienic practice and control confounding. P-value <= 0.05 was declared as statistical significance.

### 2.6. Operational definition

The students’ knowledge and practices were scored by using a scoring system adapted after reviewing different past study. Every correct answer from knowledge part was attracted 1(one) point, whereas an incorrect or don’t know the answer was attracted no (0) mark. Respondents that scored below mean under knowledge were declared as having poor knowledge; whereas those that scored above mean were declared as having good knowledge. Similarly, those students that score above mean and below mean under practice were declared as having good and poor practices respectively.

### 2.7. Ethical considerations

Ethical clearance was obtained from the IHRERC from Harar health science college and was submitted to Harari regional state of Education Bureau, the respective directors of the high schools ((SOS, Zoom in, Abadiir secondary school, Hamaresa secondary school, Harar senior school, and Bethlehem Catholic school). The purposes of the study will be explained and informed consent turned into secured from every participant. Confidentiality was maintained throughout the study. Participation in the look at became voluntarily

## 3. Result and Discussion

### 3.1 Result

#### 3.1.1 Socio-demographic characteristics

Out of 301 (100%) lady students participated in the study and the majority of students, 87% and 13% were from public and private schools respectively. from all ladies in the study mean age was 15.87 years. Among the participants 124 (41.2%) of them were in the age group of 14-15 years, 160(53.2%) of them were in the age group of 16-17 years and the rest 17(5.6%) is>= 18 years. A majority, 279(92.7%) of the participants had started their menstruation in the age range of 10-14 years.

#### 3.1.2 Knowledge Regarding menstruation and menstrual hygiene

According to the facts obtained from the participants, 184 (61.1%) study participants had good knowledge about menstrual hygiene, 117(38.9%) respondents had poor menstrual hygiene knowledge Most of the girls 275(91.4%) knew about menses before they had menarche. The dominant source of information and advice for the girls were mothers 163(54.2%). (Table).

**Table 1:** Knowledge on menstruation and menstrual hygiene management practice among female high school students in Harari region Ethiopia, 2019.

| Respondents characteristics | Frequency | Percent (%) |
| --- | --- | --- |
| **At what age girl usually get her first period?** |  |  |
| 11 - 16 years | 299 | 99.3 |
| > 16 years | 2 | 0.7 |
| **How long is the normal menstrual bleeding duration?** |  |  |
| < 2 days | 22 | 7.3 |
| 2 - 7 days | 234 | 77.7 |
| > 7 days | 28 | 9.3 |
| don't know | 17 | 5.6 |
| **What is the normal duration of menstrual cycle?** |  |  |
| < 20 days | 50 | 16.6 |
| 20 - 35 days | 200 | 66.4 |
| > 35 days | 23 | 7.6 |
| i don't know | 28 | 9.3 |
| **Do you know sanitary pads in the market?** |  |  |
| Yes | 275 | 91.4 |
| No | 26 | 8.6 |
| **Cause of menstruation** |  |  |
| Physiological Process | 264 | 87.7 |
| It Caused By Sin | 25 | 8.3 |
| Is Curse Of God | 6 | 2.0 |
| I Don't Know | 6 | 2.0 |
| **Source of bleeding during menstruation** |  |  |
| Vagina | 84 | 27.9 |
| Urinary Bladder | 16 | 5.3 |
| Uterus | 180 | 59.8 |
| I Don't Know | 21 | 7.0 |
| **Knowledge about Menstrual hygiene practice** |  |  |
| Good knowledge | 184 | 61.1 |
| Poor knowledge | 117 | 38.9 |

**Table 2:**
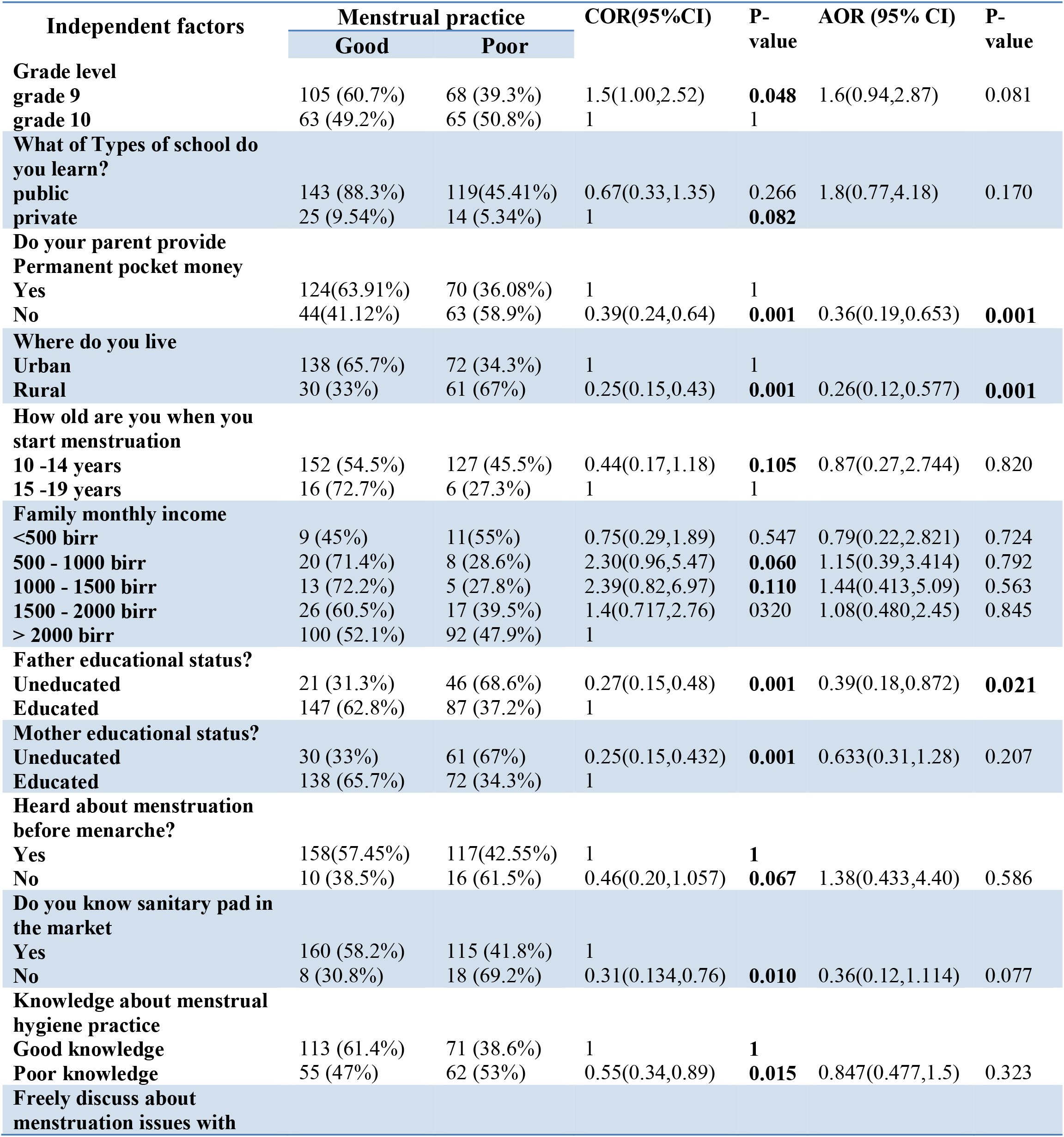
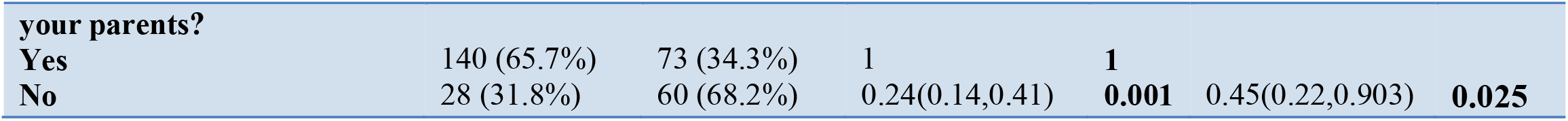
Bivariate and multivariate logistic regression analysis for Factors associated with menstrual hygiene practice of female high school students, Harari region in Eastern Ethiopia 2019.

| Independent factors | Menstrual practice |  | COR(95%CI) | P-value | AOR (95% CI) | P-value |
| --- | --- | --- | --- | --- | --- | --- |
|  | Good | Poor |  |  |  |  |
| Grade level |  |  |  |  |  |  |
| grade 9 | 105 (60.7%) | 68 (39.3%) | 1.5(1.00,2.52) | **0.048** | 1.6(0.94,2.87) | 0.081 |
| grade 10 | 63 (49.2%) | 65 (50.8%) | 1 | 1 |  |  |
| What of Types of school do you learn? |  |  |  |  |  |  |
| public | 143 (88.3%) | 119(45.41%) | 0.67(0.33,1.35) | 0.266 | 1.8(0.77,4.18) | 0.170 |
| private | 25 (9.54%) | 14 (5.34%) | 1 | **0.082** |  |  |
| Do your parent provide Permanent pocket money |  |  |  |  |  |  |
| Yes | 124(63.91%) | 70 (36.08%) | 1 | 1 |  |  |
| No | 44(41.12%) | 63 (58.9%) | 0.39(0.24,0.64) | **0.001** | 0.36(0.19,0.653) | **0.001** |
| Where do you live |  |  |  |  |  |  |
| Urban | 138 (65.7%) | 72 (34.3%) | 1 | 1 |  |  |
| Rural | 30 (33%) | 61 (67%) | 0.25(0.15,0.43) | **0.001** | 0.26(0.12,0.577) | **0.001** |
| How old are you when you start menstruation |  |  |  |  |  |  |
| 10 -14 years | 152 (54.5%) | 127 (45.5%) | 0.44(0.17,1.18) | **0.105** | 0.87(0.27,2.744) | 0.820 |
| 15 -19 years | 16 (72.7%) | 6 (27.3%) | 1 | 1 |  |  |
| Family monthly income |  |  |  |  |  |  |
| <500 birr | 9 (45%) | 11(55%) | 0.75(0.29,1.89) | 0.547 | 0.79(0.22,2.821) | 0.724 |
| 500 - 1000 birr | 20 (71.4%) | 8 (28.6%) | 2.30(0.96,5.47) | **0.060** | 1.15(0.39,3.414) | 0.792 |
| 1000 - 1500 birr | 13 (72.2%) | 5 (27.8%) | 2.39(0.82,6.97) | **0.110** | 1.44(0.413,5.09) | 0.563 |
| 1500 - 2000 birr | 26 (60.5%) | 17 (39.5%) | 1.4(0.717,2.76) | 0.320 | 1.08(0.480,2.45) | 0.845 |
| > 2000 birr | 100 (52.1%) | 92 (47.9%) | 1 |  |  |  |
| Father educational status? |  |  |  |  |  |  |
| Uneducated | 21 (31.3%) | 46 (68.6%) | 0.27(0.15,0.48) | **0.001** | 0.39(0.18,0.872) | **0.021** |
| Educated | 147 (62.8%) | 87 (37.2%) | 1 |  |  |  |
| Mother educational status? |  |  |  |  |  |  |
| Uneducated | 30 (33%) | 61 (67%) | 0.25(0.15,0.432) | **0.001** | 0.633(0.31,1.28) | 0.207 |
| Educated | 138 (65.7%) | 72 (34.3%) | 1 |  |  |  |
| Heard about menstruation before menarche? |  |  |  |  |  |  |
| Yes | 158(57.45%) | 117(42.55%) | 1 | **1** |  |  |
| No | 10 (38.5%) | 16 (61.5%) | 0.46(0.20,1.057) | **0.067** | 1.38(0.433,4.40) | 0.586 |
| Do you know sanitary pad in the market |  |  |  |  |  |  |
| Yes | 160 (58.2%) | 115 (41.8%) | 1 |  |  |  |
| No | 8 (30.8%) | 18 (69.2%) | 0.31(0.134,0.76) | **0.010** | 0.36(0.12,1.114) | 0.077 |
| Knowledge about menstrual hygiene practice |  |  |  |  |  |  |
| Good knowledge | 113 (61.4%) | 71 (38.6%) | 1 | **1** |  |  |
| Poor knowledge | 55 (47%) | 62 (53%) | 0.55(0.34,0.89) | **0.015** | 0.847(0.477,1.5) | 0.323 |
| Freely discuss about menstruation issues with |  |  |  |  |  |  |
| your parents? |  |  |  |  |  |  |
| Yes | 140 (65.7%) | 73 (34.3%) | 1 | **1** |  |  |
| No | 28 (31.8%) | 60 (68.2%) | 0.24(0.14,0.41) | **0.001** | 0.45(0.22,0.903) | **0.025** |

#### 3.1.3 Menstrual hygiene practice of respondents

**Figure 1.**
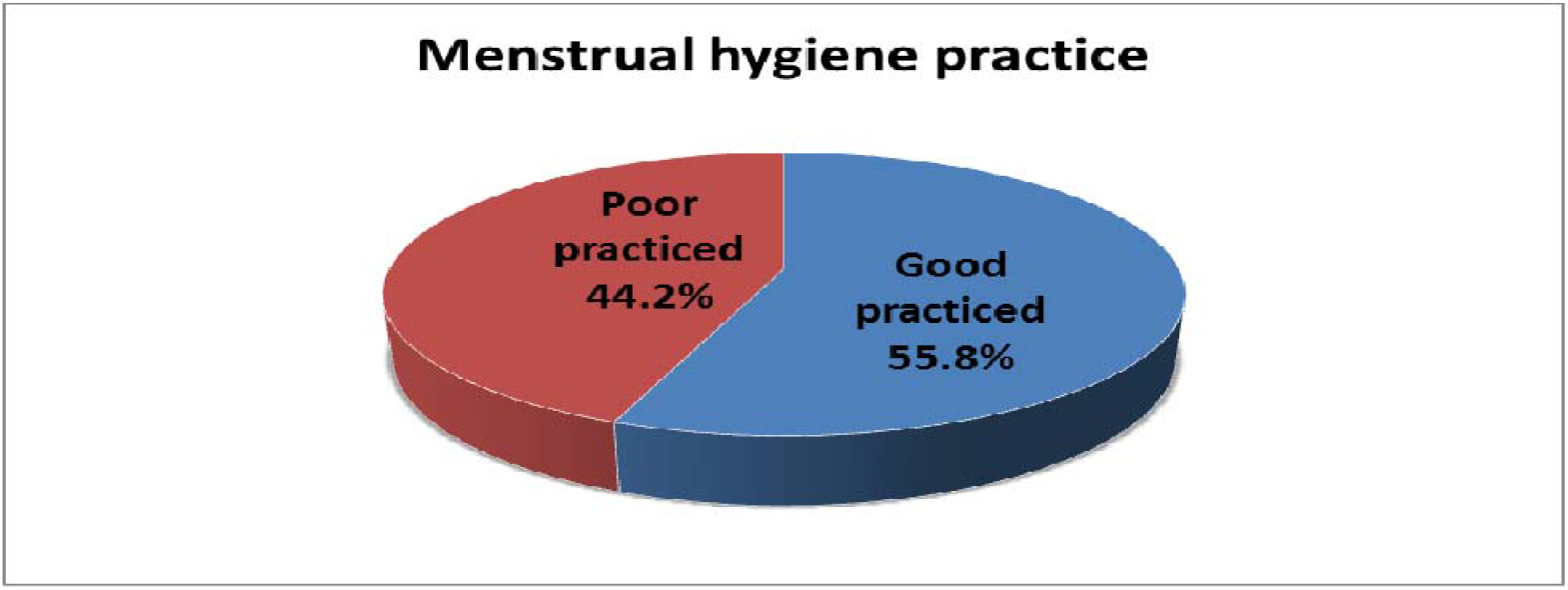
menstrual hygiene practice of high school female students in Harari region eastern Ethiopia in 2019.

According to the facts obtained from the participants, 168 (55.8%) of the respondents had a good practice and whereas 133(44.2%) practiced poor menstrual hygiene. All of the participants use sanitary pads among them 219(72.8%) use disposable sanitary materials, 35(11.6%) use reusable sanitary pads, 14(4.7%) use a disposable piece of rags and 28(9.3%) use paper/toilet paper/underwear and 5(1.7%) uses other material.

#### 3.1.4 Factors associated with menstrual hygiene practice

The students who live in the rural area (AOR 0.269: 95% CI, 0.125, 0.577) was 74% less likely to practice poor menstrual hygiene than students who live in the urban, students who have no permanent or constant pocket money from family (AOR 0.36: 95% CI, 0.309, 0.99) was 64% less likely poor menstrual hygiene practice than students who have permanent or constant pocket money from family, students who have no educated father (AOR 0.39: 95% CI, 0.18,0.872) wa 61% less likely to practice poor menstrual hygiene than students who have educated father.

### 3.2 Discussion

In this study, 184 (61.1%) study participant had good knowledge about menstrual hygiene, 117(38.9%) respondents had poor menstrual hygiene knowledge this is high when compared with the study done in Wegera District, it was found that 145(34.3%) had good knowledge regarding menses, But, it’s low when compared with similar study in Dang Nepal and eastern Ethiopia, majority 356 (87.7%) and 460 (68.5%) of female had good knowledge about menstruation, this difference may be due variation in sample size and study population. [9, 11, 12]

Out of 301 participant, 168 (55.8%) of the respondents had a good menstrual hygiene practice and whereas 133(44.2%) poor practiced during their menstrual hygiene. Similar in north-west Ethiopia and Nepal indicate out of 423 study participants 126 (29.8%) of the respondents practiced good menstrual hygiene whereas 297(70.2%) practiced poor menstrual hygiene and among 406 girls, more than half of girls (272 (67.0%)) practiced good menstrual hygiene whereas 33.0% practiced poor menstrual hygiene respectively. This difference might be the difference in students’ knowledge level regarding menstrual hygiene practice and socio-cultural difference. [9, 13]

In this study among 301 study participants, 276(91.7%) were heard about menses before menarche, A similar study done in dang Nepal, northeast and western Ethiopia, indicate, the majority of the girls 345 (84.9%), 478 (86.75%), and 247 (73%) heard about menses before menarche respectively, this almost similar findings. But, Similar study in India shows that only 42% of the ladies had knowledge about menstruation prior to onset of menarche. This difference might be due to variation in sample size and study population. [5, 7, 9, 13, 14]

According to this study, all of the study participants (301) use menstrual absorbent, other similar study conducted in northern and eastern Ethiopia, shows almost all study participants use a menstrual absorbent 100% and 614 (91.4%) respectively, this is almost similar finding. [6, 12]

This study revealed that 198(94.3%) and 77 (84.6%) residencies of the urban and rural participants respectively had awareness about menses prior to menarche. This finding is higher than a study done in Udupi Taluk, India, 83 (33.27%) and 197 (35.82%) urban and rural participants respectively, had awareness about menses prior to menarche. This difference is probably due to socio-cultural factors like talking about menstruation might be taboo in Indian mothers so the girls might not have awareness and difference in study population. [5]

In present study Among 276(91.7%) study participants were heard about menstruation before menarche and the predominant sources of information about menstruation were, 163(54.2%), 70 (23.3%), 13(4.3%), 15(5%) and 14(5%) from mothers, teacher, health personnel, TV and others respectively. In similar study conduct in Nigeria revealed 64.2% of the participants were aware of menstruation and their mothers is an essential supply of information, while buddies and TV also contributed to their information this is almost similar. [8]

This study found out the practice of good menstrual hygiene was more among students who live in the urban area (AOR, 0.269: 95% CI, 0.125, 0.577) than students who live in the rural area This finding is similar with the study done in Kano Northwestern Nigeria. It was observed that girls who live in the urban had a good practice (AOR= 2.708: 95%CI, 1.33, 3.123) of menstrual hygiene than who live in the rural area. [15]

## 4. Conclusion

According to this study, the major of high school female students in Harari region had good knowledge about menstrual hygiene practice. According to the information obtained from the study participant more than half of the female students had good menstrual hygiene practice. Good menstrual hygiene Practice was more among students who live in the urban than students who live in the rural area, good menstrual hygiene practice was more among students who have everlasting or constant pocket cash from family than students who have no permanent or regular pocket cash from family, good menstrual hygiene practice was more among students who have educated father than students who haven’t educated father and students who had Freely discuss about menses with parents had more good menstrual hygiene practice than who hadn’t Freely discuss about menses with parents.

## List of abbreviations

HBV: Hepatitis B Virus
HIV: Human Immunodeficiency virus
IHRERC: Institutional Health Research Ethics Review Committee
MHM: Menstrual hygiene management
MHP: Menstrual hygiene practices
RTI’s: Reproductive tract infections

## Declaration

### Ethics approval and consent to participate

Ethics approval letter were obtained from Harar Health Science College Research ethics review committee. Research Ethics Approval Ref/No: 2/5722/21/2/12

## Consent to participate

Written informed Consent or permission was obtained from the administrators and Parents/guardian of every selected high schools for participants under 18 years old. All study participants were informed that they have full rights to participate and withdraw from participation at any time, without any harmful events.

## Consent for publication

- Not applicable -.

## Data availability

Any time corresponding Author provide additional resource on request.

## Competing interests

The authors declare that they have no competing interests

## Funding

Self-sponsor which means there is no other funding source

## Authors’ contributions

AA and AA, participate in the study from the inception to design, acquisition of data, analysis and interpretation and MA participate interpretation and drafting of the manuscript.

## Acknowledgments

First of all, we would like to express thanks to Harar health Science College for giving us this opportunity and to thanks our advisor S/Firehiwot demise for her constructive comments, and support for the accomplishment of this research. Our deepest gratitude also goes to honored instructors Asefa Tola, Arif Hussein, Siraj Adem and Abdu Omar for their moral and material support during our study and finally we would like to thanks for those schools allowed to conduct study.

## Notes

### Competing Interest Statement

The authors have declared no competing interest.

### Funding Statement

their is no grant or funding source for this research that means it's self sponsored by both authors

### Summary of Updates

updated due to: Other Authors added based on their contribution for the manuscript, on drafting and interpritation on result.

